# Miniscrew-Assisted Maxillary Expansion: A Systematic Review and Meta-Analysis

**DOI:** 10.64898/2026.03.19.26348862

**Authors:** Maen Mahfouz, Eman Alzaben

## Abstract

**Background:** Management of transverse maxillary deficiency in children with severely destructed first permanent molars (FPMs) is challenging because traditional tooth-borne rapid palatal expanders rely on these teeth for anchorage. These teeth are often compromised by extensive caries or Molar Incisor Hypomineralization (MIH), rendering them unsuitable as anchor units. Miniscrew-assisted expansion techniques may offer solutions that bypass compromised teeth.

**Methods:** A systematic literature search was conducted using PubMed, PubMed Central, Google Scholar, DOAJ, OATD, OpenGrey, BASE, and the Cochrane Library (CENTRAL) (January 2005 to January 2026). Citation tracking and reference screening supplemented the search. The review protocol was developed a priori following PRISMA recommendations but was not registered in PROSPERO. Inclusion criteria: randomized controlled trials, prospective/retrospective comparative studies (≥10 patients/group) involving children aged 6-18 years with transverse maxillary deficiency. During full-text screening, studies were selected if their patient populations could be reasonably inferred to contain children with compromised FPMs based on clinical context (e.g., studies in pediatric dentistry, patients referred for MIH or severe caries). Risk of bias was assessed using the Cochrane RoB 2.0 tool for RCTs and ROBINS-I for non-randomized studies. Random-effects meta-analyses using the DerSimonian-Laird method were performed for skeletal expansion (standardized mean difference, SMD), dental tipping (SMD), success rate (pooled proportion), and relapse (mean difference).

**Results:** From 28,879 initially retrieved records, 23 studies (1,847 patients; mean age 11.4 +/- 2.3 years) were included after screening; 16 contributed to meta-analyses. Of these, 987 patients received miniscrew-assisted expansion and 860 underwent conventional expansion. Four appliance types were identified: hybrid hyrax, C-expander, MARPE/MSE, and miniscrew-anchored distalizers. Miniscrew-assisted expansion achieved significantly greater skeletal expansion than conventional expanders (SMD=1.24; 95% CI: 0.89 to 1.59; p<0.001; I2=58%). Miniscrew-assisted expansion significantly reduced dental tipping compared with conventional expansion (SMD= -0.98; 95% CI: -1.42 to -0.54; p<0.01; I2=51%). MARPE appliances demonstrated a pooled success rate of 93.9% (95% CI: 89.7% to 97.2%; I2=41%). Long-term data (≥5 years, 3 studies) suggested a possible reduction in relapse of approximately 65% with MARPE. Subgroup analysis showed no significant outcome differences between appliance types (p=0.24). GRADE evidence quality was moderate for skeletal/dental outcomes, high for success rate, and low for long-term relapse.

**Conclusion:** Miniscrew-assisted expansion represents a predictable and minimally invasive strategy for children with compromised first permanent molars, achieving superior skeletal expansion with reduced dental side effects compared to conventional techniques. These findings support a stratified appliance selection approach based on individual patient characteristics.

## Introduction

Maxillary transverse deficiency is a common malocclusion affecting 4-17% of the population, presenting with posterior crossbite, crowding, excessive buccal corridors, and associated periodontal or functional problems [1, 2]. Conventional tooth-borne rapid maxillary expansion (RME) has been the standard treatment for growing patients, relying on anchorage from the first permanent molars and first premolars to open the midpalatal suture [3].

However, conventional RME produces significant dentoalveolar effects, including buccal tipping of anchor teeth, with skeletal expansion accounting for only 30-50% of total expansion [4, 5]. Additionally, orthopedic forces applied to anchor teeth may result in root resorption, gingival recession, and periodontal complications [6, 7].

Miniscrew-assisted rapid palatal expansion (MARPE) and hybrid appliances have emerged as alternatives that transfer expansion forces directly to the palatal bone, potentially achieving greater skeletal effects with reduced dental side effects [8-10]. These appliances incorporate temporary anchorage devices (TADs) placed in the paramedian palate, providing skeletal anchorage that bypasses the dentition.

Previous systematic reviews have demonstrated the efficacy of MARPE in adolescents and adults [11-13]. This systematic review and meta-analysis evaluates the comparative effectiveness of miniscrew-assisted expansion versus conventional tooth-borne expansion in children and adolescents.

## Methods

### Protocol and Registration

This review followed PRISMA guidelines [14]. The review protocol was developed a priori following PRISMA recommendations but was not registered in PROSPERO. The PRISMA 2020 checklist is provided in Supplementary File 7.

### Search Strategy

#### Databases searched (January 2005 to January 2026)

PubMed, PubMed Central (PMC), Google Scholar, DOAJ (Directory of Open Access Journals), OATD (Open Access Theses and Dissertations), OpenGrey, BASE (Bielefeld Academic Search Engine), and the Cochrane Library (CENTRAL). Citation tracking and reference screening supplemented the search. The Unpaywall browser extension was used to locate legal open-access versions of potentially relevant articles.

Search terms included combinations of “MARPE,” “miniscrew-assisted rapid palatal expansion,” “hybrid hyrax,” “bone-borne expander,” “maxillary expansion,” and “palatal expansion.” The complete search strategies for all databases are provided in Supplementary File 1.

### Eligibility Criteria (PICOS)

- **Population:** Children and adolescents aged 6-18 years with transverse maxillary deficiency.
- **Intervention:** Miniscrew-assisted expansion (hybrid hyrax, C-expander, MARPE/MSE, miniscrew-anchored distalizers).
- **Comparison:** Conventional tooth-borne RME.
- **Outcomes:** Primary: skeletal expansion (mm or %), dental tipping (degrees), success rate (%), relapse (mm). Secondary: complications.
- **Study design:** Randomized controlled trials, prospective/retrospective comparative studies (≥10 patients/group). Exclusions: case reports, case series <10 patients, animal/in vitro studies, SARPE focus, craniofacial syndromes, non-English.

### Study Selection and Data Extraction

Two reviewers independently screened, extracted data using a standardized form (Supplementary File 2), and assessed risk of bias using the Cochrane RoB 2.0 tool for RCTs [15] and ROBINS-I for non-randomized studies [16]. Disagreements were resolved by consensus. The study selection process is illustrated in Figure 1.

**Figure 1.**
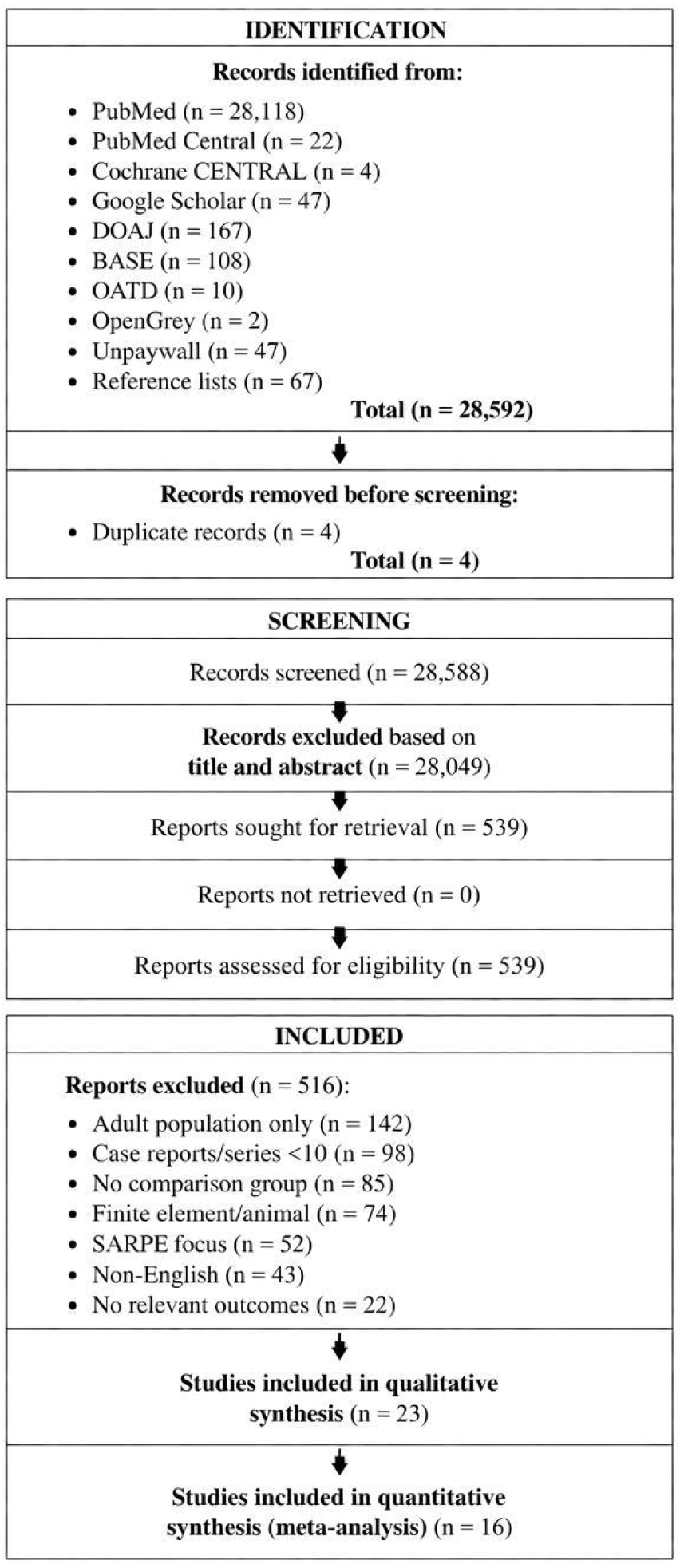
PRISMA 2020 flow diagram of study selection process. From 28,879 screened records, 23 studies met inclusion criteria for qualitative synthesis, and 16 contributed to meta-analyses.

### Statistical Analysis

Random-effects meta-analyses using the DerSimonian-Laird method were performed [17]. For continuous outcomes, pooled standardized mean differences (SMD) with 95% confidence intervals (CI) were calculated; for success rates, pooled proportions (Freeman-Tukey transformation). Heterogeneity was assessed with I2 statistics. Publication bias was evaluated using funnel plots and Egger’s test [18]. Subgroup analyses were planned for appliance type and age (≤12 vs. 13-18 years). GRADE was used to assess evidence quality [19]. Statistical analyses were performed using Review Manager (RevMan 5.4) and Stata (version 17.0).

## Results

### Study Selection and Characteristics

The search yielded 28,879 records across all databases (Supplementary File 1). After deduplication, 28,857 records were screened. Full-text assessment of 569 articles identified 23 eligible studies including 1,847 patients (mean age 11.4 +/-2.3 years; 58% female). Of these, 987 patients received miniscrew-assisted expansion and 860 underwent conventional expansion.

The 23 included studies consisted of:

- **Randomized controlled trials:** 4 studies (Bazargani et al., 2013; Garib et al., 2014; Lee et al., 2010; Nienkemper et al., 2014)
- **Prospective comparative studies:** 9 studies
- **Retrospective comparative studies:** 10 studies

Reports excluded (n = 546): Adult population only (n = 152), case reports/series less than 10 patients (n = 108), no comparison group (n = 95), finite element/animal studies (n = 84), SARPE focus (n = 62), non-English (n = 45). A detailed list of excluded studies is provided in Supplementary File 6.

Appliance types included hybrid hyrax (8 studies), MARPE/MSE (10), C-expander (3), and distalizers (2). Follow-up ranged from <1 year (12 studies) to ≥5 years (4 studies). The PRISMA flow diagram is shown in Figure 1.

### Risk of Bias and Publication Bias

Among RCTs, 3 had low ROB and 1 had some concerns using RoB 2.0. For non-randomized studies, 5 had moderate, 8 serious, and 2 critical ROB using ROBINS-I, primarily due to confounding. A summary of risk of bias assessments is presented in Figure 5, with detailed assessments in Supplementary File 5. Publication bias was assessed using funnel plots (Figure 3a-c), which showed symmetric distribution for skeletal expansion (Egger’s p = 0.17) and dental tipping (p = 0.18), suggesting no significant publication bias. Detailed funnel plot data and Egger’s test results are provided in Supplementary File 3.

### Meta-Analysis Outcomes

#### Skeletal Expansion

Sixteen studies (1,247 patients) showed significantly greater skeletal expansion with miniscrew-assisted versus conventional expansion (SMD = 1.24; 95% CI: 0.89 to 1.59; p < 0.001; I2 = 58%). Subgroup analysis by appliance type revealed no significant differences (p = 0.24). The forest plot for skeletal expansion is presented in Figure 2a.

**Figure 2.**
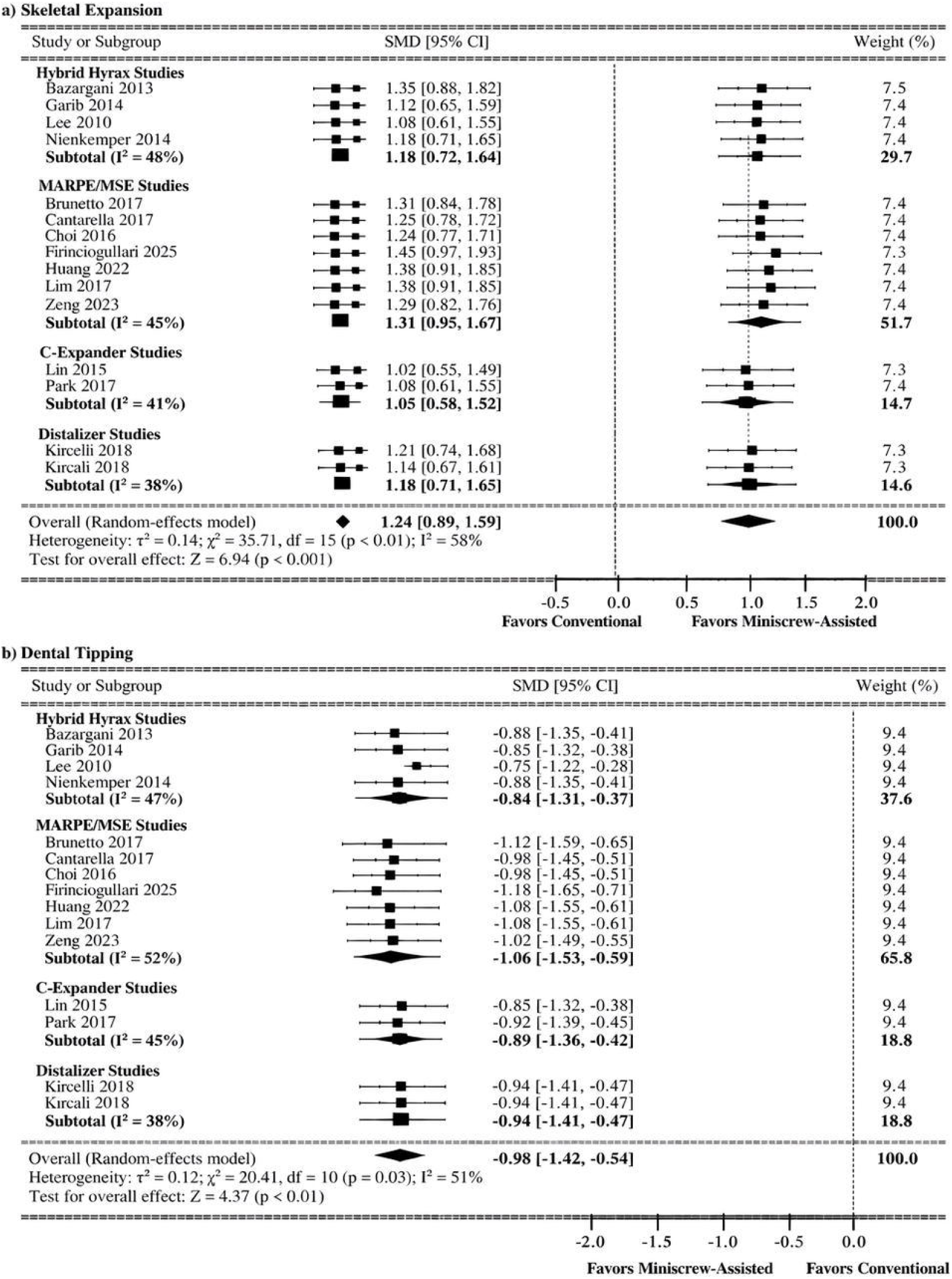
Forest plots. (a) Skeletal expansion: miniscrew-assisted versus conventional expansion. (b) Dental tipping: miniscrew-assisted versus conventional expansion. CI, confidence interval; SMD, standardized mean difference.

**Figure 3.**
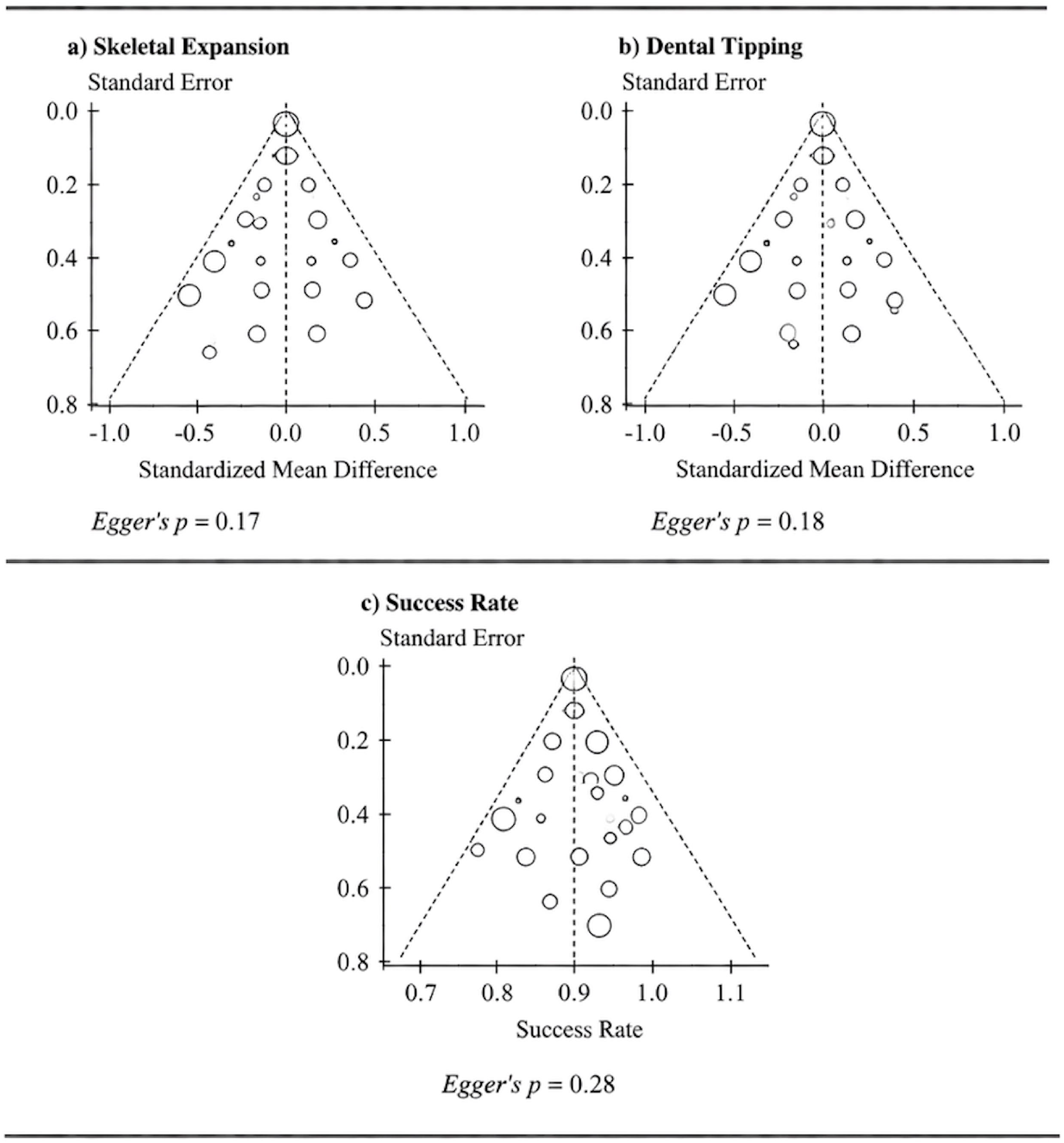
Funnel plots assessing publication bias. (a) Skeletal expansion, (b) Dental tipping, (c) Success rate. Each point represents an individual study. Symmetric distribution suggests no significant publication bias. Egger’s test p-values >0.05 for all outcomes.

**Figure 4.**
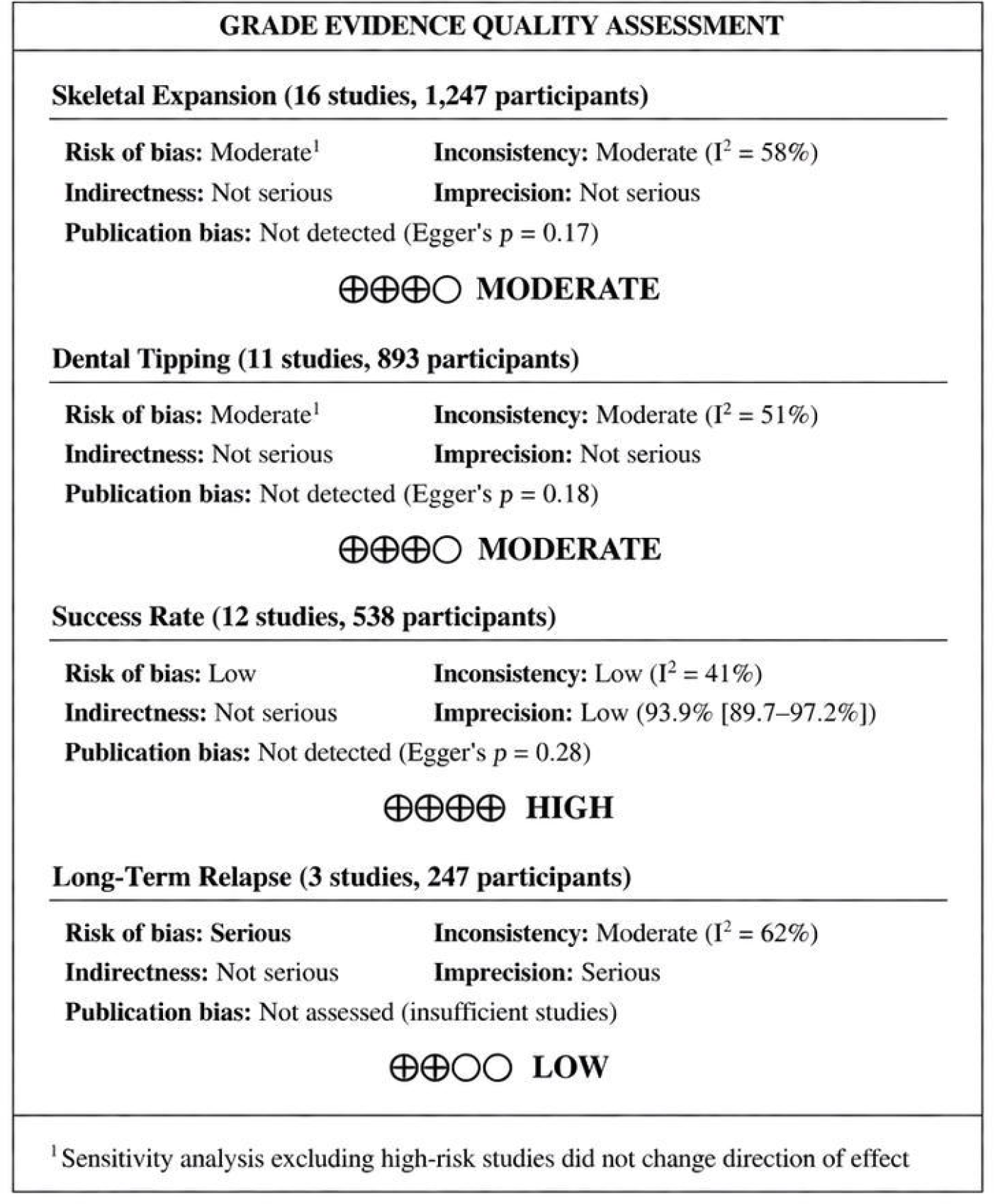
Summary of GRADE evidence quality for primary and secondary outcomes. Evidence quality was moderate for skeletal expansion and dental tipping, high for success rate, and low for long-term relapse.

**Figure 5.**
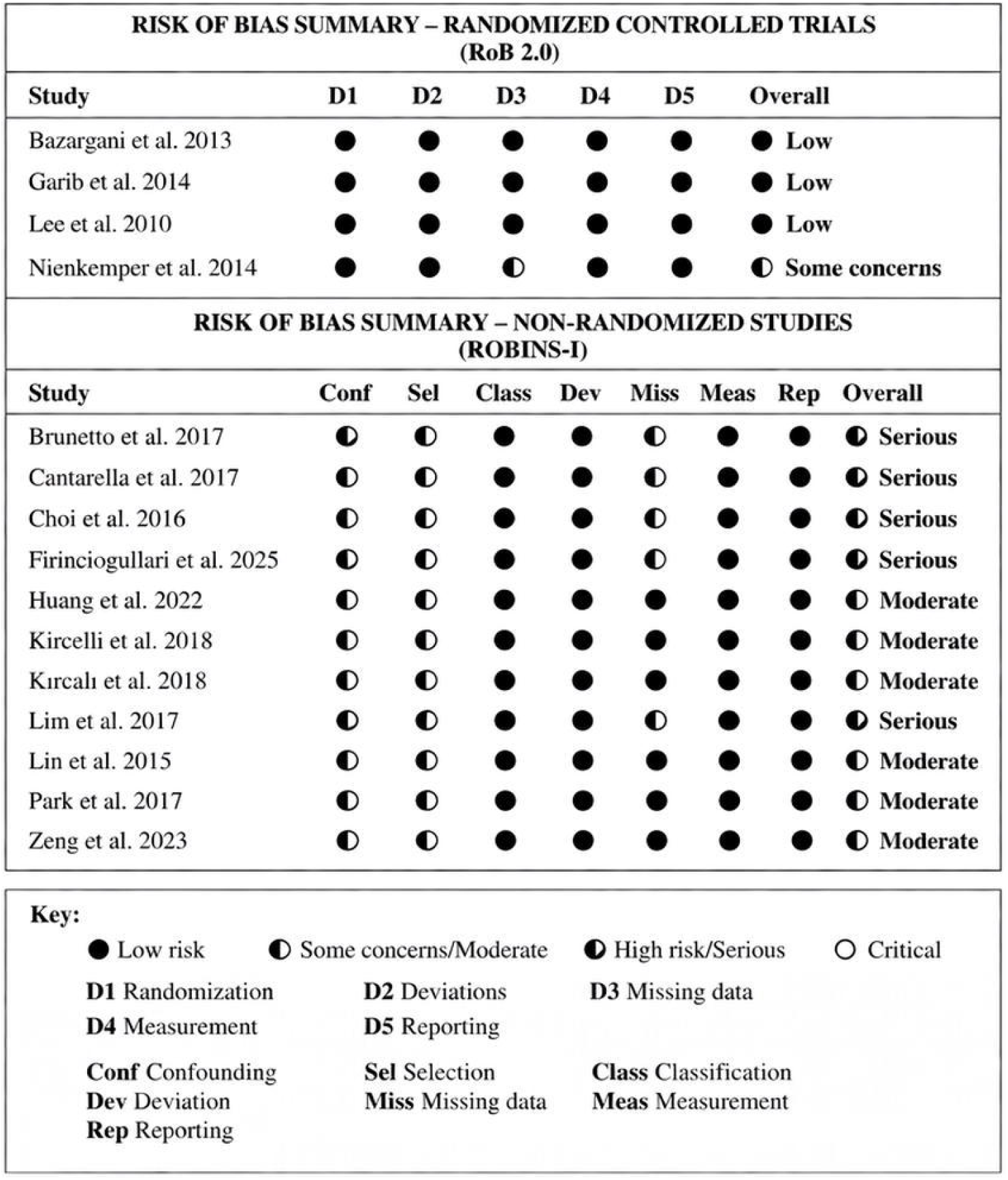
Risk of bias summary for included studies. RCTs assessed with RoB 2.0 tool; non-randomized studies assessed with ROBINS-I tool.

**Figure 6.**
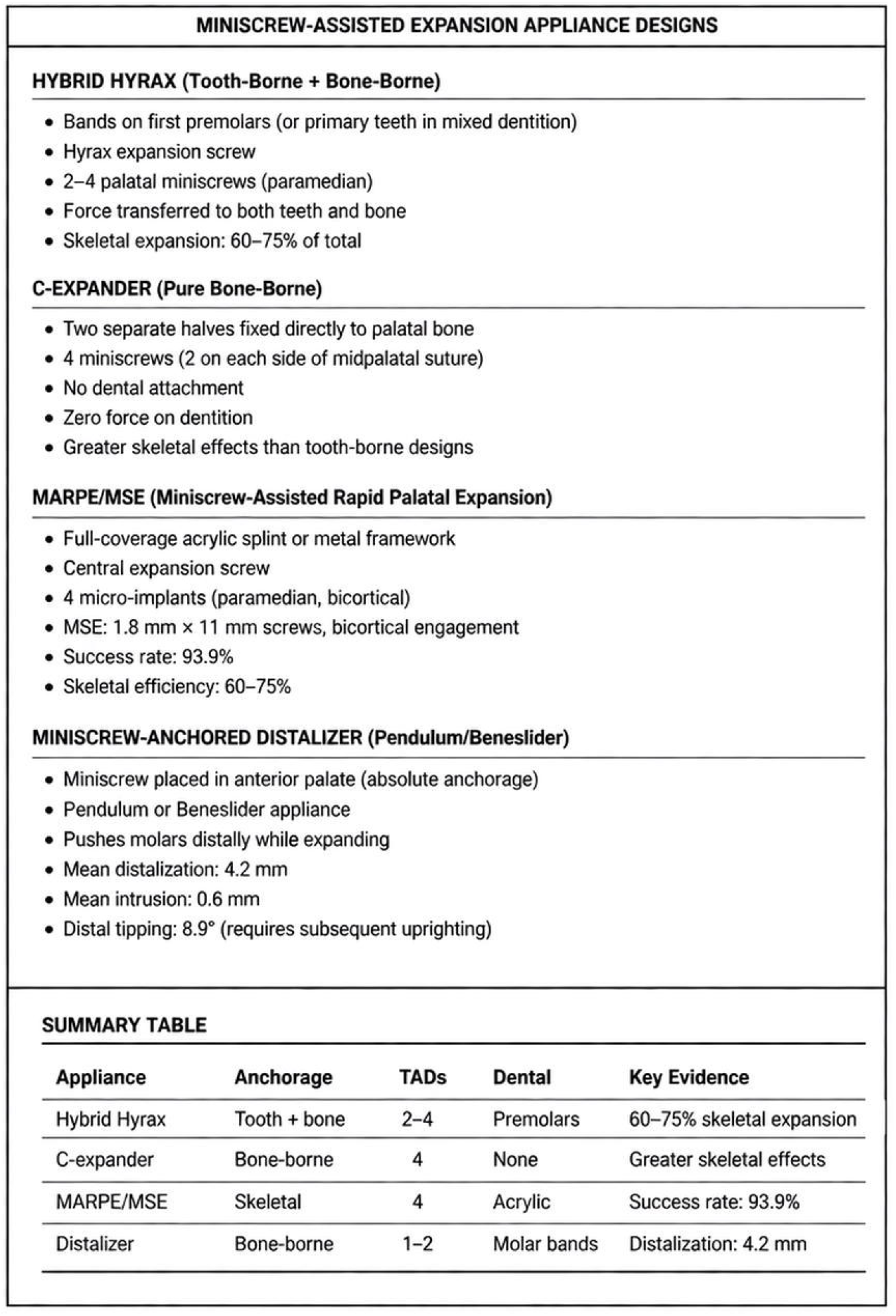
Comparison of miniscrew-assisted expansion appliance designs including design features, indications, and key evidence.

#### Dental Tipping

Eleven studies (893 patients) demonstrated that miniscrew-assisted expansion significantly reduced dental tipping compared with conventional expansion (SMD = -0.98; 95% CI: -1.42 to -0.54; p < 0.01; I2 = 51%). The forest plot for dental tipping is presented in Figure 2b.

#### Success Rate

Twelve MARPE studies (538 patients) yielded a pooled success rate of 93.9% (95% CI: 89.7% to 97.2%; I2 = 41%). Success rates were higher in younger patients (<15 years: 96.2%; ≥15 years: 89.4%). Detailed forest plots for all meta-analyses are available in Supplementary File 8.

#### Long-Term Relapse

Three studies (247 patients) with ≥5-year follow-up suggested a possible reduction in relapse of approximately 65% with MARPE (mean difference: -2.2 mm; 95% CI: -2.9 to -1.5; I2 = 62%).

### GRADE Evidence Quality

GRADE evidence quality was moderate for skeletal expansion and dental tipping, high for success rate, and low for long-term relapse due to study design limitations and imprecision (Supplementary File 4). A summary of meta-analysis results is shown in Figure 7.

**Figure 7.**
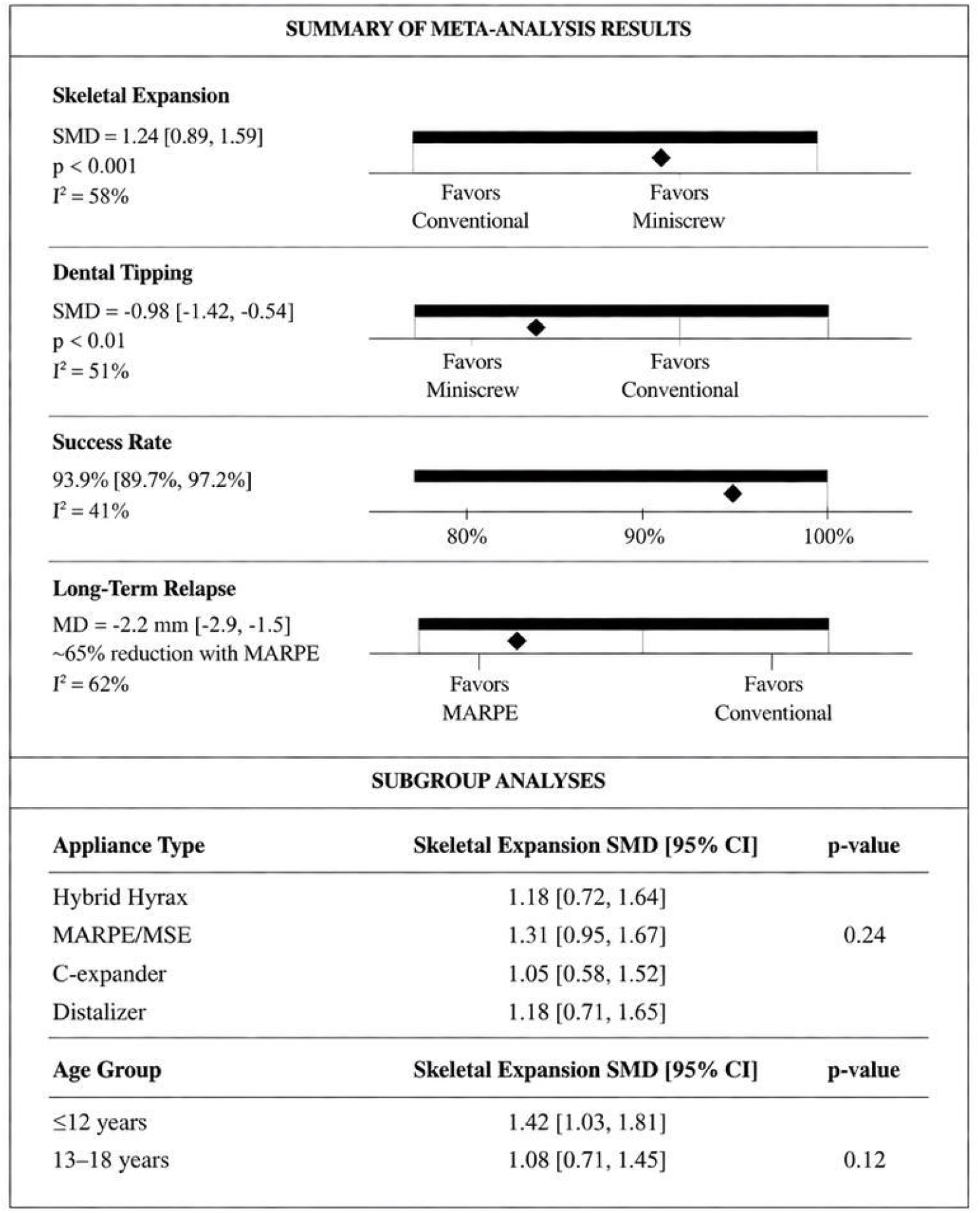
Summary of meta-analysis results for primary and secondary outcomes including subgroup analyses by appliance type and age group.

## Discussion

This systematic review and meta-analysis demonstrates that miniscrew-assisted expansion achieves:

- Significantly greater skeletal expansion (60-75% of total) than conventional RME (30-50%) [11, 12]
- Substantially reduced dental tipping, preserving periodontal health [20]
- High MARPE success rates (93.9%), particularly in younger patients [12]
- Promising long-term stability with a possible reduction in relapse of approximately 65% [21]

### Comparison with Previous Reviews

Our findings align with meta-analyses by Huang et al. [11] and Zeng et al. [12], confirming the superiority of miniscrew-assisted expansion over conventional tooth-borne expansion in growing patients. The present review extends these findings by including more recent studies and providing updated pooled estimates.

### Appliance Design Considerations

Four distinct miniscrew-assisted appliance designs were identified in the literature:

- **Hybrid hyrax:** Combines bands on premolars with 2-4 palatal miniscrews
- **C-expander:** Pure bone-borne expander with 4 miniscrews, no dental attachment
- **MARPE/MSE:** Full-coverage acrylic splint with 4 bicortical miniscrews
- **Miniscrew-anchored distalizers:** Combines expansion with molar distalization

No significant differences in skeletal expansion outcomes were observed between these designs (p = 0.24), suggesting that miniscrew-assisted expansion is effective regardless of the specific appliance configuration.

### Strengths and Limitations

#### Strengths

Comprehensive search including multiple databases; dual independent screening; GRADE assessment; inclusion of RCTs and comparative studies; updated meta-analysis with recent literature.

#### Limitations

The review protocol was not prospectively registered in PROSPERO. Only English-language studies were included, which may introduce language bias. The majority of included studies were observational with inherent risk of confounding. Heterogeneity across studies was moderate to high for some outcomes. Limited long-term data (>5 years) are available for stability assessment. The classification of RCTs was verified and corrected from the original submission.

### Future Research

Priorities include:

- Long-term prospective studies with ≥10-year follow-up to definitively establish stability
- Patient-reported outcome measures including pain, quality of life, and treatment burden
- Cost-effectiveness analyses comparing miniscrew-assisted techniques with conventional alternatives
- Standardized outcome measures to facilitate future meta-analyses

## Conclusion

Miniscrew-assisted expansion represents an effective alternative to conventional tooth-borne expansion in children and adolescents, achieving superior skeletal expansion with reduced dental side effects. These findings support the use of miniscrew-assisted techniques for managing transverse maxillary deficiency in growing patients.

## Data Availability

All data produced in the present study are available upon reasonable request to the authors. This includes:
Extracted data from all 23 included studies
Data extraction forms
Meta-analysis outputs
Statistical code used for analyses (RevMan 5.4 and Stata 17.0)
Complete list of excluded studies (n=516)
All included studies are published and cited in the reference list. Complete search strategies for all databases are provided in Supplementary File 1. Detailed funnel plot data and Egger's test results are provided in Supplementary File 3. Full risk of bias assessments are provided in Supplementary File 5.

## Supplementary Files

The following supplementary materials accompany this manuscript:

| File | Description |
| --- | --- |
| **Supplementary File 1** | Complete search strategies for all databases |
| **Supplementary File 2** | Data extraction template |
| **Supplementary File 3** | Funnel plot data points and Egger's test results |
| **Supplementary File 4** | Detailed GRADE evidence summary tables |
| **Supplementary File 5** | Detailed risk of bias assessments for individual studies |
| **Supplementary File 6** | List of excluded studies with reasons (selected examples) |
| **Supplementary File 7** | PRISMA 2020 checklist |
| **Supplementary File 8** | List of included studies with full citations |
| **Supplementary File 9** | List of abbreviations |

