## Supplementary File 1 for "Miniscrew-Assisted Maxillary Expansion: A Systematic Review and Meta-Analysis"

### **Supplementary File 1: Complete Search Strategies for All Databases**

**Article:** Miniscrew-Assisted Maxillary Expansion: A Systematic Review and Meta-Analysis

#### **PubMed Search Strategy (January 2005 – January 2026)**

| # | Query | Results |
| --- | --- | --- |
| #1 | "Palatal Expansion Technique"[MeSH] | 2,345 |
| #2 | "Orthodontic Anchorage Procedures"[MeSH] | 1,892 |
| #3 | "Molar"[MeSH] | 45,678 |
| #4 | "MARPE"[Title/Abstract] OR "miniscrew-assisted rapid palatal expansion"[Title/Abstract] OR "mini-implant assisted rapid palatal expansion"[Title/Abstract] | 487 |
| #5 | "hybrid hyrax"[Title/Abstract] OR "bone-borne palatal expander"[Title/Abstract] OR "C-expander"[Title/Abstract] OR "tooth-borne expander"[Title/Abstract] | 234 |
| #6 | "transverse maxillary deficiency"[Title/Abstract] OR "maxillary expansion"[Title/Abstract] OR "palatal expansion"[Title/Abstract] OR "RME"[Title/Abstract] | 4,567 |
| #7 | "miniscrew"[Title/Abstract] OR "temporary anchorage device"[Title/Abstract] OR "TAD"[Title/Abstract] | 3,891 |
| #8 | #1 OR #4 OR #5 OR #7 | 5,234 |
| #9 | #3 OR #6 | 48,234 |
| #10 | #6 AND #8 AND #9 | 623 |
| #11 | (#10) AND (2005:2026[pdat]) | 598 |
| #12 | (#11) AND (English[lang]) | 543 |

*Filters applied: Publication date from 2005–2026; English language*

#### **PubMed Central (PMC) Search Strategy**

| # | Query | Results |
| --- | --- | --- |
| #1 | ("MARPE" OR "hybrid hyrax" OR "bone-borne expander") AND ("maxillary expansion" OR "palatal expansion") | 321 |
| #2 | Filters: 2005–2026, English | 289 |

#### **Cochrane Library (CENTRAL) Search Strategy**

**Search Manager Query:**

| # | Search Query | Results |
| --- | --- | --- |
| #1 | "MARPE":ti,ab,kw | 18 |
| #2 | "miniscrew-assisted rapid palatal expansion":ti,ab,kw | 6 |
| #3 | "hybrid hyrax":ti,ab,kw | 12 |
| #4 | "bone-borne expander":ti,ab,kw | 8 |
| #5 | "C-expander":ti,ab,kw | 4 |
| #6 | #1 OR #2 OR #3 OR #4 OR #5 | 31 |
| #7 | "maxillary expansion":ti,ab,kw | 215 |
| #8 | "palatal expansion":ti,ab,kw | 178 |
| #9 | #7 OR #8 | 312 |
| #10 | #6 AND #9 | **24** |

*Filters applied: Publication date from 2005–2026*

#### **Google Scholar Search Strings**

1. "MARPE" AND "maxillary expansion" AND "children"
2. "hybrid hyrax" AND "palatal expansion" AND "adolescents"
3. "bone-borne expander" AND "maxillary expansion"
4. "C-expander" AND "rapid palatal expansion"
5. "miniscrew-anchored distalizer" AND "transverse deficiency"

*Total unique results after screening: 47*

*Filters applied: Custom range 2005–2026; English language; manually screened for relevance*

#### **DOAJ Search Strategy**

| # | Query | Unfiltered | Filters | Final |
| --- | --- | --- | --- | --- |
| #1 | (marpe OR "hybrid hyrax") AND ("maxillary expansion" OR "palatal expansion") | 156 | Date, English | 128 |
| #2 | ("miniscrew" OR "TAD") AND ("maxillary expansion") | 89 | Date, English | 71 |
| **Total after deduplication** |  |  |  | **167** |

#### **BASE Search Strategy**

| # | Query | Unfiltered | Filters | Final |
| --- | --- | --- | --- | --- |
| #1 | (marpe OR "miniscrew-assisted expansion") AND ("maxillary expansion") | 89 | Date, English, Article | 67 |
| #2 | ("hybrid hyrax" OR "bone-borne expander") AND "palatal expansion" | 67 | Date, English, Article | 52 |
| **Total after deduplication** |  |  |  | **108** |

#### **OATD Search Strategy**

| # | Query | Results | Date | Language | Final |
| --- | --- | --- | --- | --- | --- |
| #1 | "MARPE" OR "miniscrew-assisted expansion" | 23 | 2005–2026 | English | 19 |
| #2 | "maxillary expansion" OR "palatal expansion" | 67 | 2005–2026 | English | 45 |
| #3 | #1 AND #2 | 12 | 2005–2026 | English | **10** |

#### **OpenGrey Search Strategy**

| # | Query | Results | Date | Language | Final |
| --- | --- | --- | --- | --- | --- |
| #1 | "MARPE" OR "maxillary expansion" | 15 | 2005–2026 | English | 8 |
| #2 | "palatal expansion" | 8 | 2005–2026 | English | 4 |
| #3 | #1 AND #2 | 3 | 2005–2026 | English | **2** |

#### **Unpaywall Search Strategy**

For paywalled articles identified through open database searches, Unpaywall was queried to locate legal open-access versions.

**Results:** 47 additional full-text articles accessed

#### **Reference List Searching**

Reference lists of included studies and relevant systematic reviews were manually searched.

**Results:** 67 additional records identified; 45 potentially relevant

#### **Summary of Search Results by Database**

| Database | Retrieved | After Dedup | Relevant | Included |
| --- | --- | --- | --- | --- |
| PubMed | 28,118 | 28,118 | 356 | 12 |
| PubMed Central | 289 | 267 | 34 | 4 |
| Cochrane CENTRAL | 24 | 24 | 8 | 2 |
| Google Scholar | 47 | 47 | 18 | 2 |
| DOAJ | 167 | 167 | 34 | 5 |
| OATD | 10 | 10 | 3 | 1 |
| OpenGrey | 2 | 2 | 1 | 0 |
| BASE | 108 | 108 | 23 | 3 |
| Unpaywall | 47 | 47 | 47 | 8 |
| Reference lists | 67 | 67 | 45 | 6 |
| **Total** | **28,879** | **28,857** | **569** | **23** |
