## Supplementary File 2 for "Miniscrew-Assisted Maxillary Expansion: A Systematic Review and Meta-Analysis"

### **Supplementary File 2: Data Extraction Template**

**Article:** Miniscrew-Assisted Maxillary Expansion: A Systematic Review and Meta-Analysis

| Field | Description |
| --- | --- |
| **Study Information** |  |
| Study ID | [First author, year] |
| Country | [Country] |
| Study design | RCT / Prospective / Retrospective |
| Sample size (total) | [n] |
| Sample size (intervention) | [n] |
| Sample size (control) | [n] |
| Follow-up duration | [time] |
| **Population** |  |
| Age range (years) | [min–max] |
| Mean age (SD) | [mean +/- SD] |
| Sex (F/M) | [n / n] |
| **Intervention** |  |
| Appliance type | Hybrid / C-expander / MARPE / Distalizer |
| Number of miniscrews | [n] |
| Miniscrew dimensions | [diameter x length] |
| Insertion site | Paramedian / Other |
| Expansion protocol | [activations/day, duration] |
| Retention period | [time] |
| **Comparator** |  |
| Comparator type | Conventional RME |
| Appliance details | [description] |
| **Outcomes** |  |
| Skeletal expansion (mm) | [mean +/- SD] |
| Skeletal expansion (%) | [mean +/- SD] |
| Dental tipping (degrees) | [mean +/- SD] |
| Success rate (%) | [n/N, %] |
| Relapse (mm) | [mean +/- SD] |
| Complications | [description, n] |
| **Risk of Bias** |  |
| Overall risk | Low / Moderate / Serious / Critical |
