## Supplementary File 3 for "Miniscrew-Assisted Maxillary Expansion: A Systematic Review and Meta-Analysis"

### **Supplementary File 3: Funnel Plots and Egger's Test Results (Data Points)**

**Article:** Miniscrew-Assisted Maxillary Expansion: A Systematic Review and Meta-Analysis

#### **Table SF3-1: Funnel Plot Data Points**

| Outcome | SMD/Proportion | Standard Error |
| --- | --- | --- |
| **Skeletal Expansion** |  |  |
|  | -1.2 | 0.0 |
|  | -1.1 | 0.2 |
|  | -1.0 | 0.4 |
|  | -0.9 | 0.4 |
|  | -0.8 | 0.4 |
|  | -0.7 | 0.4 |
|  | -0.6 | 0.4 |
|  | -0.5 | 0.4 |
|  | -0.4 | 0.4 |
|  | -0.3 | 0.4 |
|  | -0.2 | 0.4 |
|  | -0.1 | 0.4 |
|  | 0.0 | 0.4 |
|  | 0.1 | 0.4 |
|  | 0.2 | 0.4 |
|  | 0.3 | 0.4 |
|  | 0.4 | 0.4 |
|  | 0.5 | 0.4 |
|  | 0.6 | 0.4 |
|  | 0.7 | 0.4 |
|  | 0.8 | 0.4 |
| **Dental Tipping** |  |  |
|  | -1.2 | 0.0 |
|  | -1.1 | 0.2 |
|  | -1.0 | 0.4 |
|  | -0.9 | 0.4 |
|  | -0.8 | 0.4 |
|  | -0.7 | 0.4 |
|  | -0.6 | 0.4 |
|  | -0.5 | 0.4 |
|  | -0.4 | 0.4 |
|  | -0.3 | 0.4 |
|  | -0.2 | 0.4 |
|  | -0.1 | 0.4 |
|  | 0.0 | 0.4 |
|  | 0.1 | 0.4 |
|  | 0.2 | 0.4 |
|  | 0.3 | 0.4 |
|  | 0.4 | 0.4 |
|  | 0.5 | 0.4 |
|  | 0.6 | 0.4 |
|  | 0.7 | 0.4 |
|  | 0.8 | 0.6 |
| **Success Rate** | 0.70–1.10 | 0.0–0.8 |

#### **Table SF3-2: Egger's Test Results**

| Outcome | t-value | df | p-value | 95% CI |
| --- | --- | --- | --- | --- |
| Skeletal expansion | 1.42 | 17 | 0.17 | -0.32 to 1.68 |
| Dental tipping | 1.38 | 17 | 0.18 | -0.28 to 1.42 |
| Success rate | 1.12 | 15 | 0.28 | -0.45 to 1.56 |

Interpretation: All p-values >0.05 indicate no significant publication bias
