## Supplementary File 4 for "Miniscrew-Assisted Maxillary Expansion: A Systematic Review and Meta-Analysis"

### **Supplementary File 4: Detailed GRADE Evidence Summary Tables**

**Article:** Miniscrew-Assisted Maxillary Expansion: A Systematic Review and Meta-Analysis

#### **Table SF4-1: GRADE Assessment for Skeletal Expansion**

| Domain | Assessment | Explanation |
| --- | --- | --- |
| **Number of studies** | 16 | 1,247 participants |
| **Study design** | Mixed (RCTs + observational) | 4 RCTs, 12 observational |
| **Risk of bias** | Moderate¹ | Majority from observational studies |
| **Inconsistency** | Moderate | I² = 58% |
| **Indirectness** | Not serious | Direct population/intervention/outcome |
| **Imprecision** | Not serious | Narrow confidence intervals |
| **Publication bias** | Not detected | Egger's p = 0.17 |
| **Overall certainty** | ⨁⨁⨁◯ MODERATE | - |

¹ Sensitivity analysis excluding high-risk studies did not change direction

#### **Table SF4-2: GRADE Assessment for Dental Tipping**

| Domain | Assessment | Explanation |
| --- | --- | --- |
| **Number of studies** | 11 | 893 participants |
| **Study design** | Mixed | 3 RCTs, 8 observational |
| **Risk of bias** | Moderate¹ | Majority from observational studies |
| **Inconsistency** | Moderate | I² = 51% |
| **Indirectness** | Not serious | Direct population/intervention/outcome |
| **Imprecision** | Not serious | Narrow confidence intervals |
| **Publication bias** | Not detected | Egger's p = 0.18 |
| **Overall certainty** | ⨁⨁⨁◯ MODERATE | - |

¹ Sensitivity analysis excluding high-risk studies did not change direction

#### **Table SF4-3: GRADE Assessment for Success Rate**

| Domain | Assessment | Explanation |
| --- | --- | --- |
| **Number of studies** | 12 | 538 participants |
| **Study design** | Mixed | 3 RCTs, 9 observational |
| **Risk of bias** | Low | High consistency across studies |
| **Inconsistency** | Low | I² = 41% |
| **Indirectness** | Not serious | Direct population/intervention/outcome |
| **Imprecision** | Low | 93.9% (95% CI 89.7–97.2%) |
| **Publication bias** | Not detected | Egger's p = 0.28 |
| **Overall certainty** | ⨁⨁⨁⨁ HIGH | - |

#### **Table SF4-4: GRADE Assessment for Long-Term Relapse**

| Domain | Assessment | Explanation |
| --- | --- | --- |
| **Number of studies** | 3 | 247 participants |
| **Study design** | Observational | Retrospective cohorts |
| **Risk of bias** | Serious | Limited number of studies |
| **Inconsistency** | Moderate | I² = 62% |
| **Indirectness** | Not serious | Direct population/intervention/outcome |
| **Imprecision** | Serious | Wide confidence intervals |
| **Publication bias** | Not assessed | Insufficient studies |
| **Overall certainty** | ⨁⨁◯◯ LOW | - |
