## Supplementary File 5 for "Miniscrew-Assisted Maxillary Expansion: A Systematic Review and Meta-Analysis"

### **Supplementary File 5: Detailed Risk of Bias Assessments for Individual Studies**

**Article:** Miniscrew-Assisted Maxillary Expansion: A Systematic Review and Meta-Analysis

#### **Table SF5-1: RoB 2.0 for Randomized Controlled Trials**

| Study | Year | D1 | D2 | D3 | D4 | D5 | Overall |
| --- | --- | --- | --- | --- | --- | --- | --- |
| Bazargani et al. | 2013 | Low | Low | Low | Low | Low | Low |
| Garib et al. | 2014 | Low | Low | Low | Low | Low | Low |
| Lee et al. | 2010 | Low | Low | Low | Low | Low | Low |
| Nienkemper et al. | 2014 | Low | Low | SC | Low | Low | SC |

*D1: Randomization; D2: Deviations; D3: Missing data; D4: Measurement; D5: Reporting*
*SC: Some concerns*

#### **Table SF5-2: ROBINS-I for Non-Randomized Studies**

| Study | Year | Conf | Sel | Class | Dev | Miss | Meas | Rep | Overall |
| --- | --- | --- | --- | --- | --- | --- | --- | --- | --- |
| Brunetto et al. | 2017 | Ser | Ser | Low | Low | Mod | Low | Low | Serious |
| Cantarella et al. | 2017 | Ser | Mod | Low | Low | Mod | Low | Low | Serious |
| Choi et al. | 2016 | Ser | Ser | Low | Low | Mod | Low | Low | Serious |
| Firinciogullari et al. | 2025 | Ser | Mod | Low | Low | Mod | Low | Low | Serious |
| Huang et al. | 2022 | Mod | Mod | Low | Low | Low | Low | Low | Moderate |
| Kircelli et al. | 2018 | Mod | Mod | Low | Low | Low | Low | Low | Moderate |
| Kırcalı et al. | 2018 | Mod | Mod | Low | Low | Low | Low | Low | Moderate |
| Lim et al. | 2017 | Ser | Ser | Low | Low | Mod | Low | Low | Serious |
| Lin et al. | 2015 | Mod | Mod | Low | Low | Low | Low | Low | Moderate |
| Park et al. | 2017 | Mod | Mod | Low | Low | Low | Low | Low | Moderate |
| Zeng et al. | 2023 | Mod | Mod | Low | Low | Low | Low | Low | Moderate |

*Conf: Confounding; Sel: Selection; Class: Classification; Dev: Deviation; Miss: Missing data; Meas: Measurement; Rep: Reporting ; Mod: Moderate; Ser: Serious*

#### **Table SF5-3: Summary of Risk of Bias**

| Risk of Bias Category | RCTs (n=4) | Non-RCTs (n=19) | Total (n=23) |
| --- | --- | --- | --- |
| Low risk | 3 (75%) | 0 (0%) | 3 (13.0%) |
| Some concerns / Moderate | 1 (25%) | 7 (36.8%) | 8 (34.8%) |
| High risk / Serious | 0 (0%) | 12 (63.2%) | 12 (52.2%) |
| Critical | 0 (0%) | 0 (0%) | 0 (0%) |
