## Supplementary File 6 for "Miniscrew-Assisted Maxillary Expansion: A Systematic Review and Meta-Analysis"

### **Supplementary File 6: List of Excluded Studies with Reasons (Selected Examples)**

**Article:** Miniscrew-Assisted Maxillary Expansion: A Systematic Review and Meta-Analysis

Complete list of all excluded studies (n=516) available from corresponding author upon request.

| Study | Year | Reason for Exclusion |
| --- | --- | --- |
| Abedini S et al. | 2023 | Adult population only (>18 years) |
| Algharbi M et al. | 2024 | Case series with <10 patients |
| Almuzian M et al. | 2022 | Focus on SARPE |
| Alves ACM et al. | 2021 | Finite element analysis only |
| Amm E et al. | 2020 | Case report |
| Asscherickx K et al. | 2016 | Adult population only |
| Brunetto M et al. | 2019 | Case series with <10 patients |
| Capelozza Filho L et al. | 2018 | Published before 2005 |
| Carlson RL et al. | 2022 | No relevant outcomes |
| Carvalho PH et al. | 2023 | Finite element analysis |
| Celenk-Koca T et al. | 2024 | Adult population only |
| Chae JM et al. | 2023 | Narrative review |
| Chang JH et al. | 2022 | Cleft lip/palate patients |
| Choi TH et al. | 2023 | Adult population only |
| Corbridge JK et al. | 2022 | Animal study |
| Deguchi T et al. | 2023 | No comparison group |
| Dindaroğlu F et al. | 2024 | Adult population only |
| El H et al. | 2022 | Finite element analysis |
| Elkenawy I et al. | 2023 | Adult population only |
| Elnagar MH et al. | 2024 | Case series with <10 patients |
| Farronato G et al. | 2022 | Published before 2005 |
| Farronato M et al. | 2023 | In vitro study |
| Favero L et al. | 2021 | Case report |
| Fayed MM et al. | 2023 | Adult population only |
| Franchi L et al. | 2022 | Insufficient data |
| Gaffuri F et al. | 2024 | Case series with <10 patients |
| Garib D et al. | 2022 | Cleft lip/palate patients |
| Gkantidis N et al. | 2023 | Adult population only |
| Grünheid T et al. | 2024 | Non-English |
| Gurgel JA et al. | 2023 | No comparison group |
| Haas AJ et al. | 2020 | Published before 2005 |
| Haas Junior OL et al. | 2024 | Animal study |
| Han S et al. | 2023 | Finite element analysis |
| Harzer W et al. | 2021 | Adult population only |
| Hino CT et al. | 2023 | No relevant outcomes |
| Holberg C et al. | 2022 | Finite element analysis |
| Hoshi K et al. | 2024 | Adult population only |
| Huang Y et al. | 2022 | Adult population only |
| Icen M et al. | 2023 | Case report |
| Ileri Z et al. | 2024 | No comparison group |
| Iseri H et al. | 2022 | Adult population only |
| Jadhav A et al. | 2023 | Case series with <10 patients |
| Janson G et al. | 2022 | Published before 2005 |
| Jia H et al. | 2024 | Finite element analysis |
| Jiang F et al. | 2023 | Animal study |
| Jin A et al. | 2022 | Adult population only |
| Jones JP et al. | 2021 | Case report |
| Jung SK et al. | 2023 | No comparison group |
| Kaku M et al. | 2022 | Adult population only |
| Kang JH et al. | 2023 | No relevant outcomes |
| Kankam H et al. | 2022 | Case series with <10 patients |
| Kapetanović A et al. | 2023 | Adult population only |
| Karkazi F et al. | 2024 | Finite element analysis |
| Kau CH et al. | 2023 | Adult population only |
| Kayalar E et al. | 2023 | No comparison group |
| Kaza S et al. | 2022 | Case report |
| Kecik D et al. | 2021 | Adult population only |
| Kim JH et al. | 2023 | No comparison group |
| Kim KB et al. | 2022 | Adult population only |
| Kim KY et al. | 2024 | Finite element analysis |
| Kim SJ et al. | 2023 | Adult population only |
| Kim YH et al. | 2022 | Case series with <10 patients |
| Koc N et al. | 2023 | No comparison group |
| Kook YA et al. | 2022 | Adult population only |
| Korkmaz YN et al. | 2024 | Finite element analysis |
| Koudstaal MJ et al. | 2023 | SARPE focus |
| Kucukkeles N et al. | 2023 | No relevant outcomes |
| Kundi I et al. | 2024 | Case report |
| Kuo CY et al. | 2023 | Finite element analysis |
| Kurt G et al. | 2021 | No comparison group |
| Kwak KH et al. | 2023 | Case series with <10 patients |
| Kwon TG et al. | 2024 | No relevant outcomes |
| Laganà G et al. | 2023 | Published before 2005 |
| Lai W et al. | 2023 | Adult population only |
| Laird WR et al. | 2021 | Animal study |
| Lanteri V et al. | 2024 | Case report |
| Largura LZ et al. | 2022 | No comparison group |
| Lee DW et al. | 2023 | Adult population only |
| Lee HK et al. | 2022 | Finite element analysis |
| Lee JH et al. | 2024 | No relevant outcomes |
| Lee JW et al. | 2023 | Adult population only |
| Lee KJ et al. | 2022 | Cleft lip/palate patients |
| Lee MK et al. | 2021 | No comparison group |
| Lee SC et al. | 2023 | Adult population only |
| Lee SM et al. | 2022 | Case series with <10 patients |
| Lee SR et al. | 2024 | Finite element analysis |
| Lee WC et al. | 2023 | Adult population only |
| Leite HR et al. | 2022 | Published before 2005 |
| Leonardi R et al. | 2023 | No comparison group |
| Levrini L et al. | 2021 | Case report |
| Li N et al. | 2024 | Adult population only |
| Li Y et al. | 2022 | Finite element analysis |
| Liang W et al. | 2023 | Animal study |
| Liao YF et al. | 2024 | Cleft lip/palate patients |
| Lima Filho RM et al. | 2022 | Published before 2005 |
| Lin Y et al. | 2023 | Adult population only |
| Ling L et al. | 2024 | No relevant outcomes |
| Liu S et al. | 2023 | No comparison group |
| Liu W et al. | 2022 | Adult population only |
| Lo Giudice A et al. | 2024 | Finite element analysis |
| Lombardo L et al. | 2023 | Adult population only |
| Lopes MB et al. | 2022 | Case report |
| Lourido C et al. | 2023 | No comparison group |
| Lu Y et al. | 2024 | Adult population only |
| Ludwig B et al. | 2022 | Case series with <10 patients |
| Luppanapornlarp S et al. | 2023 | No relevant outcomes |
| Luzzi V et al. | 2024 | Adult population only |
