## Supplementary File 7 for "Miniscrew-Assisted Maxillary Expansion: A Systematic Review and Meta-Analysis"

### **Supplementary File 7: PRISMA 2020 Checklist**

**Article:** Miniscrew-Assisted Maxillary Expansion: A Systematic Review and Meta-Analysis

| Section | Item | Checklist Item | Reported |
| --- | --- | --- | --- |
| **TITLE** | 1 | Identify as systematic review | Title page |
| **ABSTRACT** | 2 | Structured summary | Abstract |
| **INTRODUCTION** | 3 | Rationale | Introduction |
|  | 4 | Objectives | Introduction |
| **METHODS** | 5 | Eligibility criteria | Methods |
|  | 6 | Information sources | Methods |
|  | 7 | Search strategy | Methods / SF1 |
|  | 8 | Selection process | Methods |
|  | 9 | Data collection | Methods / SF2 |
|  | 10 | Data items | Methods / SF2 |
|  | 11 | Risk of bias | Methods / SF5 |
|  | 12 | Effect measures | Methods |
|  | 13 | Synthesis methods | Methods |
|  | 14 | Reporting bias | Methods / SF3 |
|  | 15 | Certainty assessment | Methods / SF4 |
| **RESULTS** | 16a | Study selection | Results / Fig 1 |
|  | 16b | Excluded studies | SF6 |
|  | 17 | Study characteristics | Results |
|  | 18 | Risk of bias | Results / SF5 |
|  | 19 | Individual study results | Results |
|  | 20a | Synthesis results | Results |
|  | 20b | Heterogeneity | Results |
|  | 20c | Subgroup analyses | Results |
|  | 20d | Sensitivity analyses | Results |
|  | 21 | Reporting biases | Results / SF3 |
|  | 22 | Certainty of evidence | Results / SF4 |
| **DISCUSSION** | 23a | Interpretation | Discussion |
|  | 23b | Limitations | Discussion |
|  | 23c | Implications | Discussion |
| **OTHER** | 24a | Registration | Methods |
|  | 24b | Protocol | Methods |
|  | 24c | Amendments | N/A |
|  | 25 | Support | Title page |
|  | 26 | Competing interests | Title page |
|  | 27 | Data availability | Data Availability |
