## Supplementary File 9 for "Miniscrew-Assisted Maxillary Expansion: A Systematic Review and Meta-Analysis"

### **Supplementary File 9: List of Abbreviations**

**Article:** Miniscrew-Assisted Maxillary Expansion: A Systematic Review and Meta-Analysis

| Abbreviation | Full Form |
| --- | --- |
| BASE | Bielefeld Academic Search Engine |
| CENTRAL | Cochrane Central Register of Controlled Trials |
| CI | Confidence Interval |
| DOAJ | Directory of Open Access Journals |
| GRADE | Grading of Recommendations Assessment, Development and Evaluation |
| I² | Heterogeneity statistic |
| MARPE | Miniscrew-Assisted Rapid Palatal Expansion |
| MD | Mean Difference |
| MeSH | Medical Subject Headings |
| MSE | Maxillary Skeletal Expander |
| OATD | Open Access Theses and Dissertations |
| PICOS | Population, Intervention, Comparison, Outcomes, Study design |
| PMC | PubMed Central |
| PRISMA | Preferred Reporting Items for Systematic Reviews and Meta-Analyses |
| PROSPERO | International Prospective Register of Systematic Reviews |
| RCT | Randomized Controlled Trial |
| RME | Rapid Maxillary Expansion |
| RoB 2.0 | Risk of Bias 2.0 tool |
| ROBINS-I | Risk Of Bias In Non-randomized Studies - of Interventions |
| SARPE | Surgically Assisted Rapid Palatal Expansion |
| SD | Standard Deviation |
| SMD | Standardized Mean Difference |
| TAD | Temporary Anchorage Device |
